# The Clinical Significance of CEA, CA19-9, and CA125 in Management of Appendiceal Adenocarcinoma

**DOI:** 10.1101/2023.09.10.23295319

**Authors:** Abdelrahman Yousef, Mahmoud Yousef, Mohammad Zeineddine, Aditya More, Saikat Chowdhury, Mark Knafl, Paul Edelkamp, Ichiaki Ito, Yue Gu, Vinay Pattalachinti, Zahra Alavi Naini, Fadl Zeineddine, Jennifer Peterson, Kristin Alfaro, Wai Chin Foo, Jeff Jin, Neal Bhutiani, Victoria Higbie, Christopher Scally, Bryan Kee, Scott Kopetz, Drew Goldstein, Abhineet Uppal, Michael G. White, Beth Helmink, Keith Fournier, Kanwal Raghav, Melissa Taggart, Michael J. Overman, John Paul Shen

## Abstract

**Importance:** Serum tumor markers CEA, CA19-9, & CA125 have been useful in the management of gastrointestinal and gynecological cancers, however there is limited information regarding their utility in patients with appendiceal adenocarcinoma.

**Objective:** Assessing the association of serum tumor markers (CEA, CA19-9, and CA125) with clinical outcomes, pathologic, and molecular features in patients with appendiceal adenocarcinoma.

**Design:** This is a retrospective study with results reported in 2023. The median follow-up time was 43 months.

**Setting:** Single tertiary care comprehensive cancer center.

**Participants:** Under an approved Institutional Review Board protocol, the Palantir Foundry software system was used to query the MD Anderson internal patient database to identify patients with a diagnosis of appendiceal adenocarcinoma and at least one tumor marker measured at MD Anderson between 2016 and 2023.

**Results:** A total of 1,338 patients with appendiceal adenocarcinoma were included, with a median age of 56.5 years. The majority of the patients had metastatic disease (80.7%). CEA was elevated in more than half of the patients tested (56%), while CA19-9 and CA125 were elevated in 34% and 27%, respectively. Individually, elevation of CEA, CA19-9, or CA125 were associated with worse 5-year survival; 82% vs 95%, 84% vs 92%, and 69% vs 93% elevated vs normal for CEA, CA19-9, and CA125 respectively (all p<0.0001). Quantitative evaluation of tumor markers increased prognostic ability. Patients with highly elevated (top 10^th^ percentile) CEA, CA19-9 or CA125 had markedly worse survival with 5-year survival rates of 59%, 64%, and 57%, respectively (HR vs. normal : 9.8, 6.0, 7.6, all p<0.0001). Although metastatic tumors had higher levels of all tumor markers, when restricting survival analysis to 1080 patients with metastatic disease elevated CEA, CA19-9 or CA125 were all still associated worse survival (HR vs. normal : 3.4, 1.8, 3.9, p<0.0001 for CEA and CA125, p=0.0019 for CA19-9). Interestingly tumor grade was not associated with CEA or CA19-9 level, while CA-125 was slightly higher in high relative to low-grade tumors (18.3 vs. 15.0, p=0.0009). Multivariable analysis identified an incremental increase in the risk of death with an increase in the number of elevated tumor markers, with a 11-fold increased risk of death in patients with all three tumor markers elevated relative to those with none elevated. Mutation in *KRAS* and *GNAS* were associated with significantly higher levels of CEA and CA19-9.

**Conclusions:** These findings demonstrate the utility of measuring CEA, CA19-9, and CA125 in the management of appendiceal adenocarcinoma. Given their prognostic value, all three biomarkers should be included in the initial workup of patients diagnosed with appendiceal adenocarcinoma.

**Key Points:** *Question:* Can serum tumor markers CEA, CA19-9, or CA125 be useful in management of patients with appendiceal adenocarcinoma?

*Findings:* In this single institution retrospective cohort study, elevation of CEA, CA19-9, or CA125 were associated with significantly worse 5-year survival; 82% vs 95%, 84% vs 92%, and 69% vs 93% elevated vs normal respectively. Moreover, quantitative evaluation of tumor markers increased prognostic ability. Further analysis identified an incremental increase in the risk of death with an increase in the number of elevated tumor markers, with a 11-fold increased risk of death in patients with all three tumor markers elevated relative to those with none elevated.

*Meaning:* Given their prognostic value, all three biomarkers should be included in the initial workup of patients diagnosed with appendiceal adenocarcinoma.

## Introduction

Appendiceal adenocarcinoma (AA) is a heterogenous disease, with marked contrast in the natural history of low-grade and high-grade tumors (5-year overall survival (OS) 68% for low-grade vs. 7% for high-grade)^1–4^. Unfortunately for the majority of patients (∼74%) there is already metastatic disease at the time of diagnosis^5,6^. Metastatic spread of AA is almost exclusively limited to the peritoneal cavity, causing the clinical syndrome pseudomyxoma peritonei (PMP) in which the peritoneal surfaces and omentum are involved with diffuse gelatinous, mucinous implants^7,8^. However, PMP progression is difficult to measure with traditional cross-sectional imaging as it frequently exists as a contiguous erratically shaped area in the peritoneal cavity. In addition, current RECIST criteria do not consider mucinous/cystic disease as measurable. For these reasons, standard RECIST criteria are poorly applicable to AA^9^. Moreover, AA is a slowly progressive disease, and classically defined thresholds for determining changes in disease extent (typically ≥20% increase) may take years to occur. For these reasons, having a more dependable method to distinguish patients with AA who are more or less likely to have favorable disease outcomes would be highly advantageous for guiding patient discussions and treatment options.

Serum tumor markers (TMs) are commonly used for aid in evaluating diagnosis, prognosis, and treatment response in different types of malignancies^10,11^. Three TMs have been well established in gastrointestinal cancers: cancer embryonic antigen (CEA), cancer antigen 19-9 (CA19-9), and cancer antigen 125 (CA125)^12–14^. These TMs have been associated with metastatic dissemination of tumor cells^15–17^. While serum markers have been useful in detecting gastrointestinal tumors, there is a lack of information regarding their efficacy in patients with AA. Researchers have suggested using CEA as a potential TM based on its utility in detection and management of colon cancer^12^. However, studies conducted thus far have not established a definitive and consistent correlation between any of the TMs and AA prognosis^18–21^.

The objectives of this study were to investigate the association between serum TM levels and clinical outcomes, as well as pathologic and molecular features across the spectrum of AA. We hypothesized that elevation in any of the three TM levels would be linked to a decline in 5-year survival, independent of other factors^10,11,22–24^. We started an active collaboration with the data science firm Syntropy^25,26^ to deploy the Palantir Foundry software platform. The Foundry platform aids in the integration, analysis, extraction, and transformation of clinical data, allowing the many elements of the Electronic Health Record (EHR) to be merged into a dataset amenable to research analyses^27,28^. The Foundry platform was utilized to conduct this study.

## Materials and Methods

Under approved Institutional Review Board (IRB) protocol Lab09-0373 (PI: Dr. Scott Kopetz), the Palantir Foundry software system (Palantir, Denver, CO)^25,26^ was used to query the MD Anderson internal patient database to identify patients with a diagnosis of AA and at least one TM measured at MD Anderson between 2016 to 2023 for inclusion in this retrospective study. Data cutoff point was May 12^th^, 2023.

The value of each TM measured was categorized as normal, elevated, or highly elevated. TMs levels below the laboratory upper limit of normal (CEA ≥ 3 ng/mL, CA19-9 ≥ 35 U/mL, and CA125 ≥ 35 U/mL, from 2016 till March 2018, and CEA > 3.8 ng/mL, CA19-9 > 35 U/mL, and CA125 > 38 U/mL, from April 2018 till 2023) were defined as normal. TMs levels above the laboratory upper limit of normal were defined as elevated; while levels in the top 10^th^ percentile for each respective TM (>99.8 ng/mL for CEA, >338.6 U/mL for CA19-9, and >99.0 U/mL for CA125) were defined as highly elevated. For patients with more than one measurement for each TM, the highest measurement was considered for analysis. Other clinical information collected through the Foundry platform included patient demographics, histopathology, tumor grade, surgical history, and mutational profiles for patients who had next generation sequencing (NGS) performed at MD Anderson. Histologic classification and grade were collected from patients’ pathology records. Pathologic diagnosis was determined by a team of expert pathologists using a three tier classification^29^. Well-differentiated and well to moderately-differentiated tumors were considered low-grade tumors, while moderately-differentiated, moderately to poorly-differentiated, and poorly-differentiated tumors were considered high-grade tumors. Low-grade tumors lacked high cellularity, invasive implants, or significant cytologic atypia. High-grade tumors exhibited invasive implants, cytologic atypia, and signet ring cells. Overall survival (OS) was defined as the time from initial diagnosis with appendiceal cancer until death.

Survival analyses were performed using the Kaplan-Meier (KM) method with log-rank test. Univariate and multivariate Cox-proportional hazards (CPH) regression analyses were performed to assess relationships between clinical factors (serum TMs levels, demographics, patient and disease characteristics) and patient outcomes (PFS and OS). KM and CPH analyses modeled TMs individually (Model A) as well as in a single aggregate variable indicating the number of elevated TMs (Model B). In both models, gender, race, smoking status, alcohol use, age at diagnosis, tumor histology, tumor grade, and tumor stage were also considered. All variables with p<0.05 on univariate analysis were included in the multivariate analysis. Differences in tumor histopathological types between patients with elevated versus normal TMs levels were assessed using Fischer’s exact test. Differences in TMs levels between tumor grade and gender were assessed using Mann-Whitney U test, while differences in TMs levels between tumor histopathology was assessed using Kruskal-Wallis test as TMs levels didn’t follow Gaussian distribution. Correlation between the different TMs were assessed using Spearman’s correlation. Frequency of mutations among patients with elevated versus normal TMs levels was assessed using Fisher’s exact test. All statistical analysis was performed using Graph Pad Prism software version 9.0.0 for Windows (GraphPad Software, San Diego, California USA, www.graphpad.com) and RStudio (RStudio team (2020) RStudio: Integrated Development for R. RStudio, PBC, Boston, MA, http://www.rstudio.com/).

## Results

### Patient Characteristics

Between 2016 to 2023, a total of 1,338 patients who had a diagnosis of AA and at least one TM (CEA, CA19-9, or CA125) measured at MD Anderson were identified **(Fig 1)**. The median follow-up times from AA diagnosis were 42.6 months. The median age of the patients was 56.5 years (**Table 1)**. The study population included slightly more females (56.3%), and the majority of patients self-reported white race (79.7%). Most of the patients in our study had stage IV metastatic disease (80.7%). More than half (52.4%) of the patients had mucinous tumors; the remainder had colonic (9.8%), goblet cell (7.0%), signet ring cell (16.7%), or a mix of signet ring and goblet cell (11.1%) histopathology. Overall, 57.2% of patients had high grade tumors (moderately or poorly differentiated) **(Table 1; Fig S1)**.

**Figure 1.**
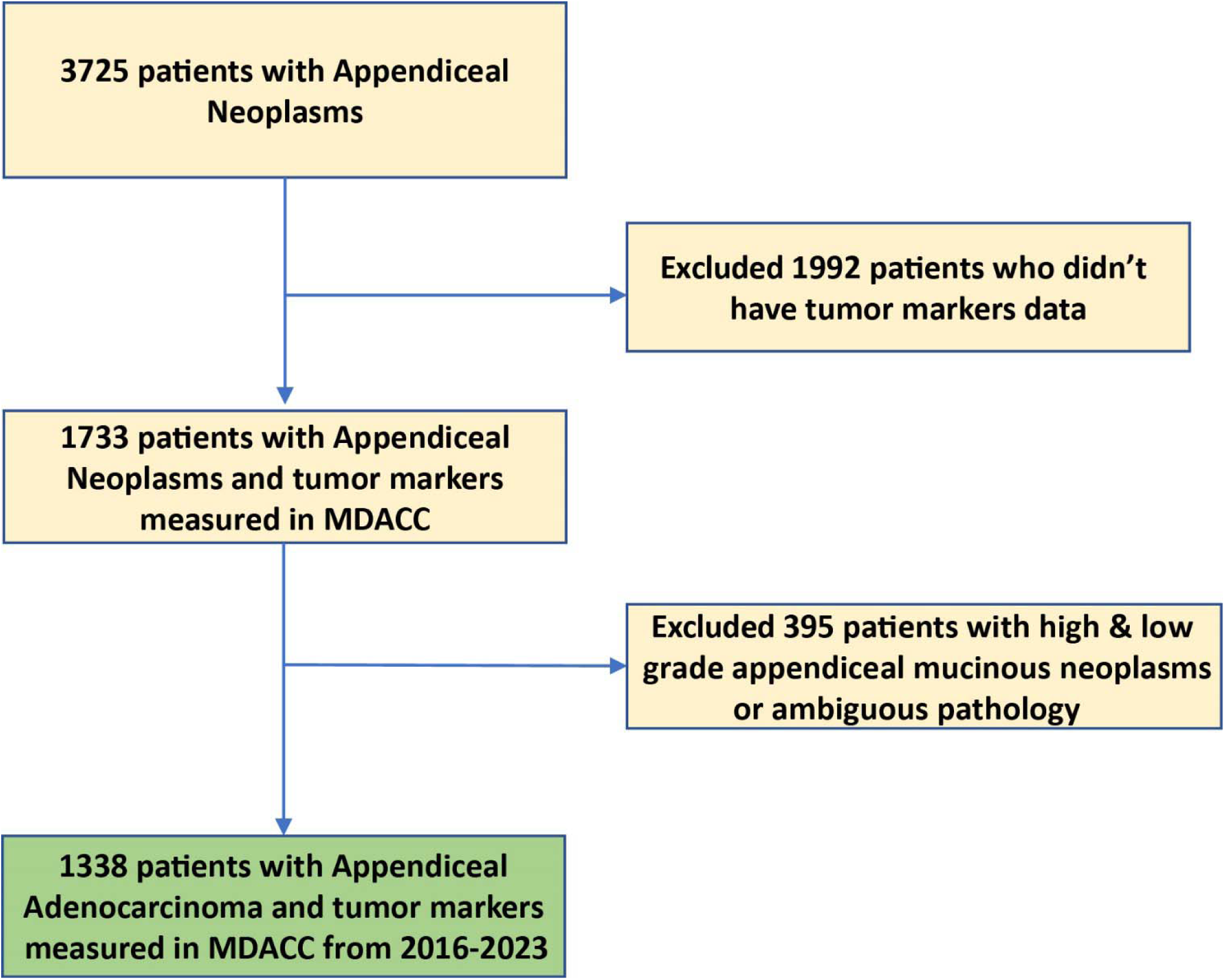
Flowchart diagram showing cohort patients selection. Abbreviations include MDACC (MD Anderson Cancer Center).

**Table 1.**
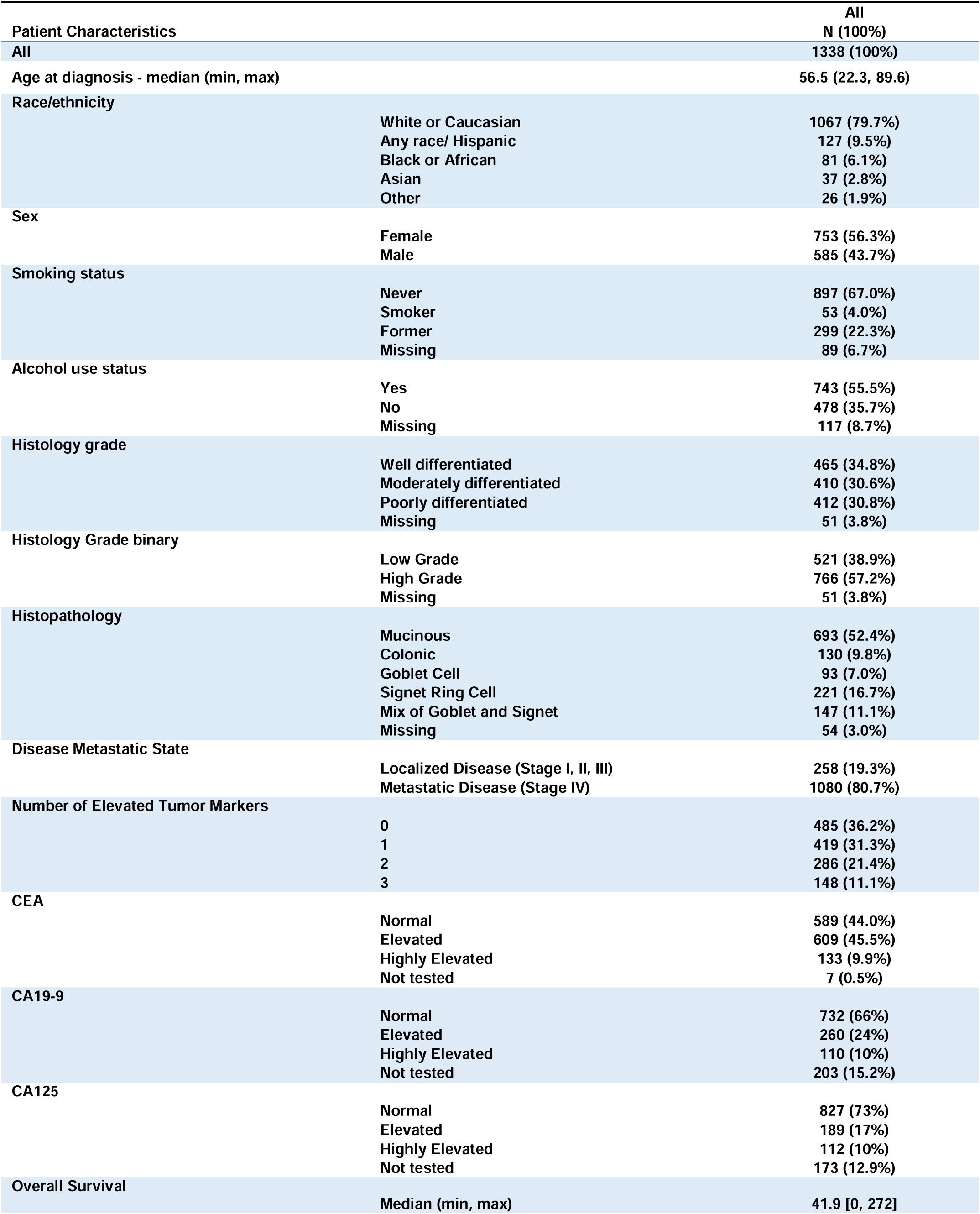
Patient Characteristics of our cohort.

| Patient Characteristics |  | All<br>N (100%) |
| --- | --- | --- |
| All |  | 1338 (100%) |
| Age at diagnosis - median (min, max) |  | 56.5 (22.3, 89.6) |
| Race/ethnicity | White or Caucasian | 1067 (79.7%) |
|  | Any race/ Hispanic | 127 (9.5%) |
|  | Black or African | 81 (6.1%) |
|  | Asian | 37 (2.8%) |
|  | Other | 26 (1.9%) |
| Sex | Female | 753 (56.3%) |
|  | Male | 585 (43.7%) |
| Smoking status | Never | 897 (67.0%) |
|  | Smoker | 53 (4.0%) |
|  | Former | 299 (22.3%) |
|  | Missing | 89 (6.7%) |
| Alcohol use status | Yes | 743 (55.5%) |
|  | No | 478 (35.7%) |
|  | Missing | 117 (8.7%) |
| Histology grade | Well differentiated | 465 (34.8%) |
|  | Moderately differentiated | 410 (30.6%) |
|  | Poorly differentiated | 412 (30.8%) |
|  | Missing | 51 (3.8%) |
| Histology Grade binary | Low Grade | 521 (38.9%) |
|  | High Grade | 766 (57.2%) |
|  | Missing | 51 (3.8%) |
| Histopathology | Mucinous | 693 (52.4%) |
|  | Colonic | 130 (9.8%) |
|  | Goblet Cell | 93 (7.0%) |
|  | Signet Ring Cell | 221 (16.7%) |
|  | Mix of Goblet and Signet | 147 (11.1%) |
|  | Missing | 54 (3.0%) |
| Disease Metastatic State | Localized Disease (Stage I, II, III) | 258 (19.3%) |
|  | Metastatic Disease (Stage IV) | 1080 (80.7%) |
| Number of Elevated Tumor Markers | 0 | 485 (36.2%) |
|  | 1 | 419 (31.3%) |
|  | 2 | 286 (21.4%) |
|  | 3 | 148 (11.1%) |
| CEA | Normal | 589 (44.0%) |
|  | Elevated | 609 (45.5%) |
|  | Highly Elevated | 133 (9.9%) |
|  | Not tested | 7 (0.5%) |
| CA19-9 | Normal | 732 (66%) |
|  | Elevated | 260 (24%) |
|  | Highly Elevated | 110 (10%) |
|  | Not tested | 203 (15.2%) |
| CA125 | Normal | 827 (73%) |
|  | Elevated | 189 (17%) |
|  | Highly Elevated | 112 (10%) |
|  | Not tested | 173 (12.9%) |
| Overall Survival |  |  |
|  | Median (min, max) | 41.9 [0, 272] |

### Tumor Marker Assessment

CEA was the most frequently tested TM (n=1331 patients), CA19-9 and CA125 were also evaluated in the majority of patients (n=1132 and 1165, respectively) **(Table 1)**. CEA was elevated in more than half of the patients tested (56%, including highly elevated levels), while CA19-9 and CA125 were elevated in 34% and 27%, respectively (including highly elevated levels), of the patients tested; 11% of patients had elevated levels of all 3 TMs **(Table 1; Table S1; Fig 2A,B).** In aggregate 70%, 59%, and 61% of patients were tested at time of diagnosis for CEA, CA19-9, and CA125 respectively, with a trend of increased utilization of all three markers starting in 2018 **(Fig S2).**

**Figure 2.**
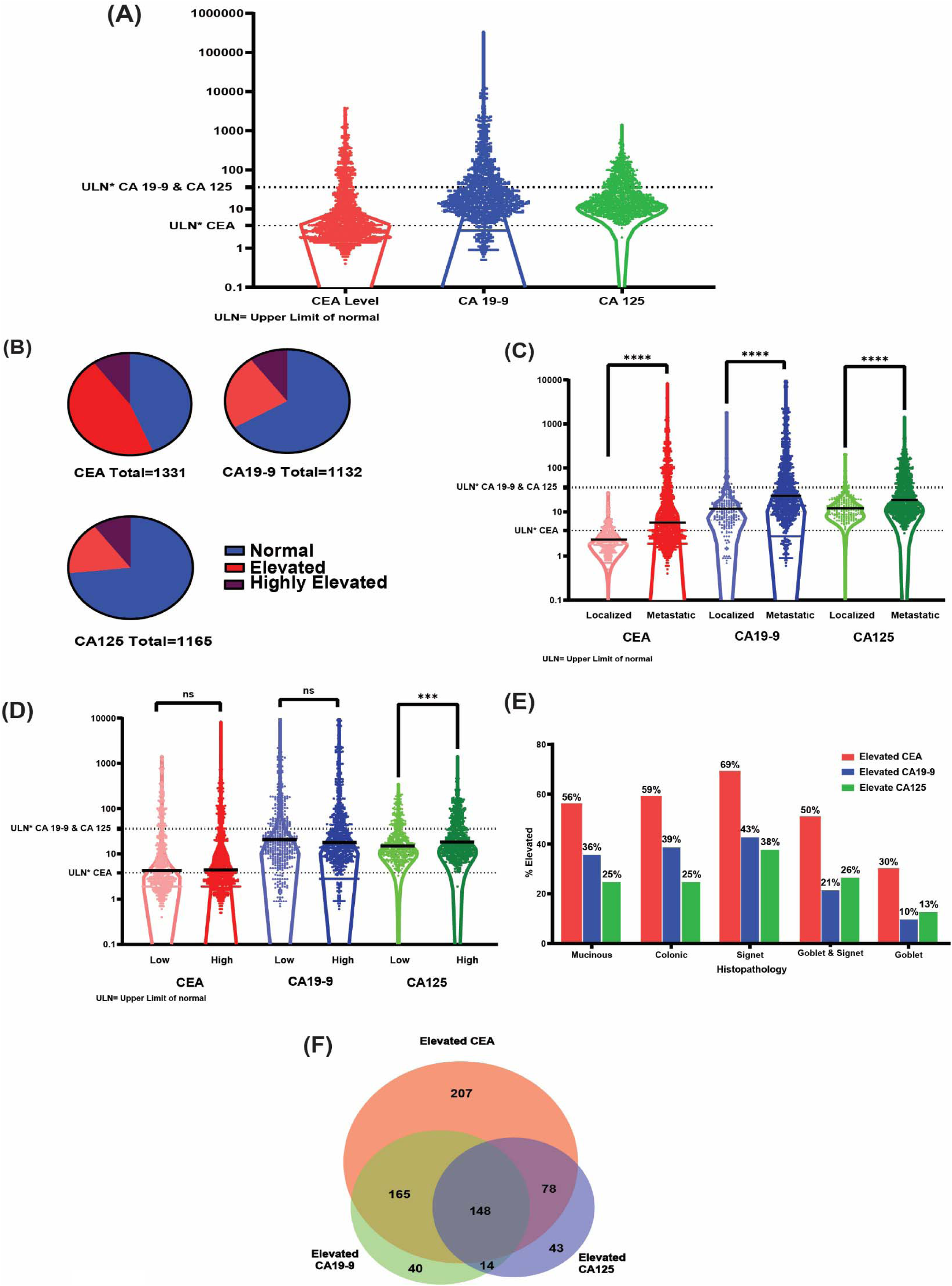
(A) Violin plot showing the distribution of all patients CEA, CA19-9, and CA125 tumor markers levels, each point represents one patient. (B) Pie charts for the three tumor markers showing the distribution of normal, elevated, and highly elevated in each. (C) Violin plot showing the distribution of all patients CEA, CA19-9, and CA125 tumor markers levels split by disease stage (metastatic vs localized), lines represent median levels (D) Violin plot showing the distribution of all patients CEA, CA19-9, and CA125 tumor markers levels split by grade. lines represent median levels (E) Bar graph showing the percentage of elevated tumor markers in different histopathological groups. (F) Venn diagram showing the overlapping of elevated (highly elevated included) levels of the three tumor markers.

CA19-9 and CA125 levels were statistically equivalent between males and females, CEA was higher in males (median of males=4.5 vs. females=4.1, p=0.024) **(Fig S3).** CEA, CA19-9, and CA125 were significantly higher in patients with metastatic disease compared to patients with localized disease (median of metastatic vs. localized= 5.8 vs. 2.2 for CEA, 23.3 vs. 11.6 for CA19-9, and 18.5 vs. 12.5 for CA125, p<0.001 for each) **(Fig 2C).** CEA and CA19-9 were statistically equivalent for high- and low-grade tumors, (p=0.40 and p=0.55, respectively), CA125 was slightly higher in high-grade tumors (median of high grade vs. low grade= 18.3 vs. 15.0, p=0.0009) **(Fig 2D; Table S2).** Comparing across histology, all three TMs were most often elevated in patients with signet ring cell histology (69, 43, and 38% for CEA, CA19-9, and CA125, p<0.001 for each) and least likely to be elevated in Goblet cell adenocarcinoma **(Fig 2E; Fig S4; Tables S3-S5)**. When measured on the same day CEA and CA19-9 levels were highly correlated (r = 0.63, p<0.0001) but there was a subset of patients with highly elevated CEA but normal CA19-9, consistent with fact that the carbohydrate CA19-9 cannot be produced in patients who genetically lack the Lewis Antigen A^30^. Both CEA and CA19-9 were correlated to CA125 but to a lesser degree(r=0.29, r=0.24, p<0.0001 for each) **(Fig S5; Table S6)**. In patients who had all the 3 markers measured (n=1,112), isolated CEA elevation was more common (18.6%) than CA19-9 (3.6%) or CA125 (3.9%), all three TM were elevated in 11.0% of patients **(Fig 2F)**^31^.

### Outcomes

KM survival analysis of OS by TM marker level (normal, elevated or highly elevated) demonstrated that CEA, CA19-9, and CA125 were all prognostic of OS (all p<0.0001) **(Fig 3A, C, E**). Compared to 5-year OS of 92-95% in the patients with normal values for each TM, 5-year with elevated CEA, CA19-9, or CA125 was 82, 84%, and 70%, respectively (HR=4.0, 95% CI 2.9-5.6, for elevated CEA, HR=2.2, 95% CI 1.4-3.4, for elevated CA19-9, and HR=4.6, 95% CI 2.7-7.8 for elevated CA125, p<0.0001 for all). Moreover, 5-year OS for those with highly elevated CEA, CA19-9, or CA125 was 61%, 66%, and 60%, respectively (HR= 9.8, 95% 5.3-18.0, for highly elevated CEA, HR=6.0, 95% CI 3.0-11.7, for highly elevated CA19-9, and HR=7.6, 95% CI 3.5-16.5 for highly elevated CA125, p<0.0001 for all). (**Fig 3G**). Given the association of metastasis with increased TM levels, the survival analysis for each TM was repeated restricting to only patients with metastatic disease (n=1080). Elevated levels of all TMs remained associated with overall survival (HR elevated vs. normal : 3.4, 1.8, 3.9, for CEA, CA19-9, and CA125, respectively, p<0.0001 for CEA and CA125, p=0.0019 for CA19-9) **(Fig 3B, D, F)** as well as highly elevated levels (HR highly elevated vs. normal : 7.4, 4.7, 6.4, p<0.0001 for all). Survival analysis for each TM was again repeated controlling for tumor grade. Elevated levels of all TM remained associated with overall survival in both low-grade and high-grade tumor subgroups (all p<0.0001) **(Fig S4)**. As a further control, analysis was restricted to those patients with TMs measured within the initial six months from the date of diagnosis (CEA, n=560, CA19-9, n=291, CA125, n=475) to allow for assessment of TMs at time of diagnosis. Again, survival analysis by TM level (normal, elevated or highly elevated) demonstrated that CEA, CA19-9, and CA125 were all prognostic of OS (HR=2.4, 95% CI 1.4-3.9, for elevated CEA, HR= 3.6, 95% 1.4-9.3, for highly elevated CEA, p=0.0006, HR=2.1, 95% CI 0.93-4.6, for elevated CA19-9, HR=2.9, 95% CI 1.1-7.9 for highly elevated CA19-9, p=0.037, HR=2.4, 95% CI 1.2-4.6 for elevated CA125, and HR=4.0, 95% CI 2.0-7.8 for highly elevated CA125, p<0.0001) (Fig S6).

**Figure 3.**
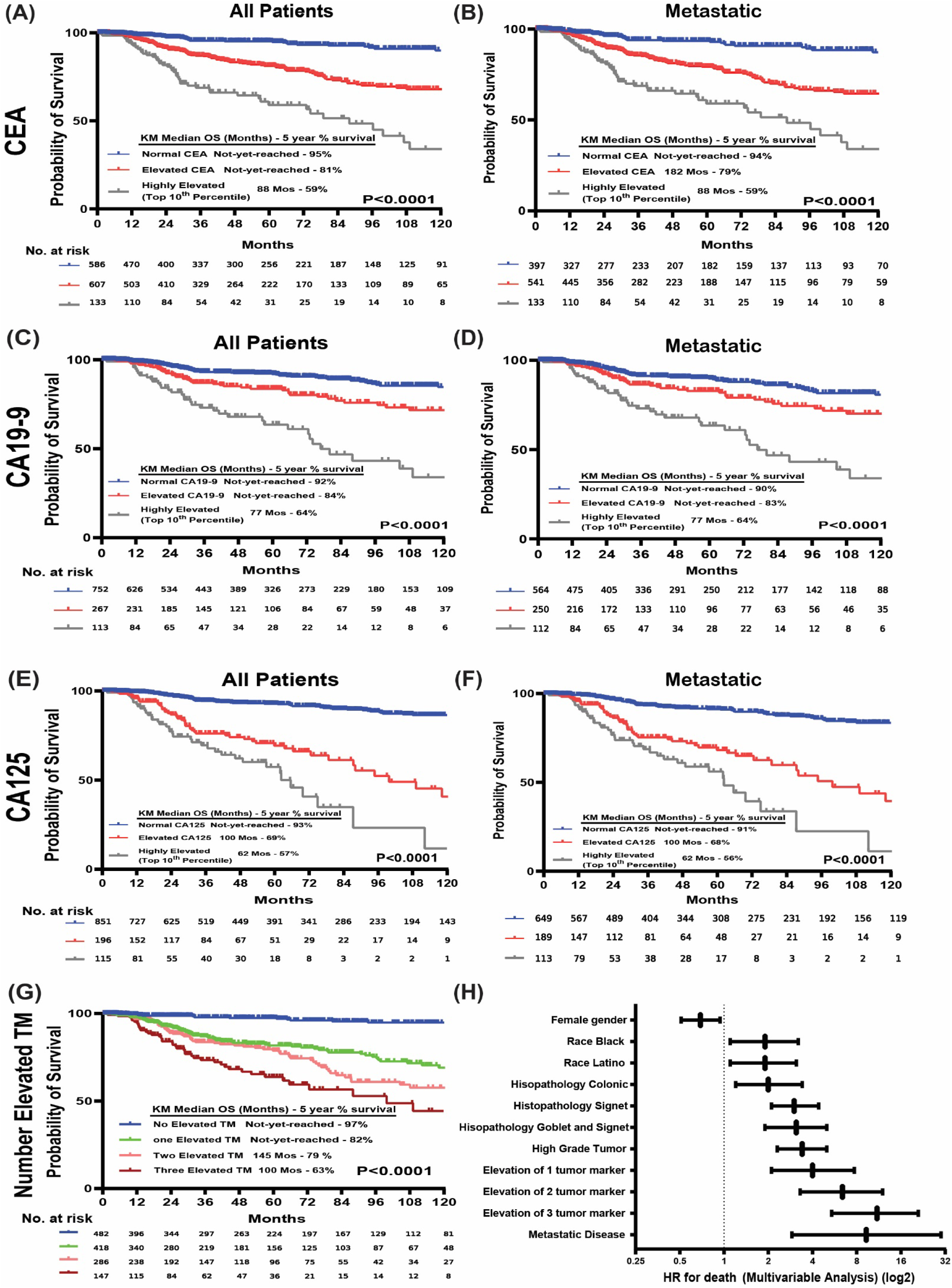
(A) KM survival plot of all patients with normal, elevated, and highly elevated levels of CEA. (B) KM survival plot of stage IV metastatic disease patients with normal, elevated, and highly elevated levels of CEA (C) KM survival plot of all patients with normal, elevated, and highly elevated levels of CA19-9. (D) KM survival plot of stage IV metastatic disease patients with normal, elevated, and highly elevated levels of CA19-9. (E) KM survival plot of all patients with normal, elevated, and highly elevated levels of CA125. (F) KM survival plot of stage IV metastatic disease patients with normal, elevated, and highly elevated levels of CA125. (F) KM survival plot of all patients with number of elevated tumor markers. (G) Forest plot for multivariable analysis showing HR for death in all patients on a log2 axis.

In multivariable analysis, After controlling for race goblet/signet histology, tumor grade, and tumor stage elevated CEA (HR=2.8, 95% CI 1.7-4.9, p=0.0001), elevated CA19-9 (HR=1.5, 95% CI 1-2.2, p=0.028), and elevated CA125 (HR=3.2, 95% CI 2.2-4.7, p<0.0001) remained significantly associated with decreased OS **(Table S7)**. Notably, when the same variables were modeled together with the number of elevated TMs, there was an incremental increase in the risk of death with an increase in the number of elevated TMs, with a 11-fold increased risk of death in patients with 3 elevated TMs relative to those with no elevated TMs (95% CI 5.4-21, p<0.0001) **(Fig 3H, Table S8).** Similar findings were observed in the subset of patients who had their TMs measured within the first 6 months from diagnosis (n=560) **(Fig S7)**

A subset of 398 tumors were also profiled with a targeted mutation panel; CEA and CA19-9 were more frequently elevated in patients with *KRAS* mutation (79% vs. 64% for CEA, p=0.003, 64% vs. 33% for CA19-9, p<0.0001, **Fig 4A**). Similarly, both CEA and CA19-9 were more frequently elevated in patients with *GNAS* mutation (89% vs. 72% for CEA, p=0.001, 68% vs. 43% for CA19-9, p<0.0001, **Fig 4B**). CEA and CA19-9 levels were higher in patients with *KRAS* mutation (median=22.0 vs. 6.0, p<0.0001 for CEA, and 80.4 vs. 18.9, p<0.0001 for CA19-9) **(Fig 4D)** and *GNAS* mutation (median=34.3 vs. 7.6, p<0.0001 for CEA, and 94.0 vs. 26.2, p=0.0002 for CA19-9) **(Fig 4E)**. *TP53* mutation was not associated with differences in TM level (**Fig 4C**). CA-125 levels were statistically equivalent regardless of mutation status for *KRAS*, *GNAS*, and *TP53*. 88 patients had elevated values for all three TMs (pan-elevated TM group) and 69 patients had normal values for all three TMs (the pan-normal TM group). Prevalence of *KRAS* and *GNAS* mutations was significantly higher in the pan-elevated group than in the pan-normal group (66% vs. 22% and 44% vs. 11% respectively, p<0.0001 for each) **(Fig 4F).** Finally, (**Fig 4G)** displays significant co-elevation between CEA and CA19-9 (OR= 9.4, 95% CI 6.8-12.9, p<0.0001), CEA and CA125 (OR= 4.9, 95% CI 3.6-6.7, p<0.0001), and CA19-9 and CA125 (OR= 4.1, 95% CI 3.1-5.4, p<0.0001). In our cohort, *KRAS* and *GNAS* were the most mutated genes (52% and 33%, respectively).

**Figure 4.**
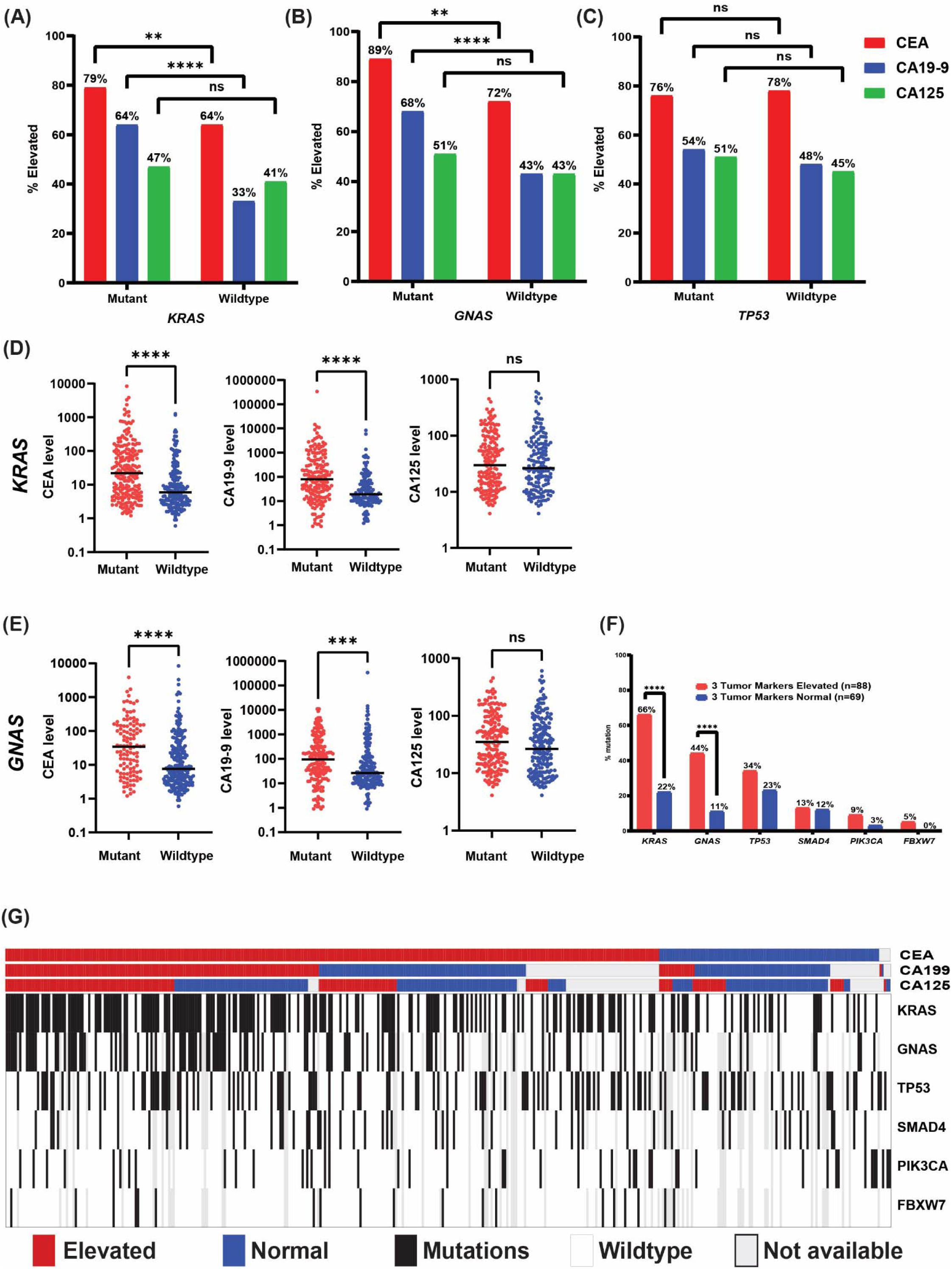
(A-C) Bar graphs showing the percentage of patients with elevated CEA (red), CA19-9 (blue), and CA125 (green) in mutant and wildtype *KRAS*, *GNAS*, and *TP53*. (D) Scattered plots for *KRAS* mutant vs wildtype with CEA, CA19-9 and CA125 levels, lines represents the median levels. (E) Scattered plots for *GNAS* mutant vs wildtype with CEA, CA19-9 and CA125 levels, lines represents the median levels. (F) Bar graph showing the prevalence of *KRAS*, *GNAS*, *TP53, SMAD4, PIK3CA, and FBXW7* (Genes that are most frequently mutated in our cohort) mutations in patients with the 3 tumor markers elevated (pan-elevated) vs. patients with normal levels of the 3 tumor markers (pan-normal). (G) Oncoplot showing the 3 tumor markers elevated levels on the left and normal levels on the right, and the mutation status of *KRAS, GNAS, TP53, SMAD4, PIK3CA*, and *FBXW7*.

## Discussion

This study represents the first comprehensive evaluation of the clinical utility of the tumor markers CEA, CA19-9, and CA125 in more than a thousand patients with appendiceal adenocarcinoma, establishing that each of the three are prognostic biomarkers. In 2018 the Chicago Consensus Working Group for the first time developed guidelines for the treatment appendiceal cancer; these endorsed the measurement of CEA, CA19-9 and CA125 in all metastatic appendiceal cancer patients^32^. However, in practice even at this NCCN designated, tertiary referral center testing of all thee tumor markers was not universal (**Fig S2**). Here we show that using the combination of CEA, CA19-9, and CA125 can stratify patients with appendiceal adenocarcinoma into groups with five-year survival ranging from 97% for those with no tumor markers elevated to 63% for those with all three markers elevated (**Figure 2G**). Our results are consistent with and expand upon multiple prior retrospective analyses in small cohorts^33,34^ and prior studies restricted to patients undergoing cytoreductive surgery^35,36^ which have suggested prognostic value of these tumor markers. In addition to demonstrating the importance of measuring all three of CEA, CA19-9 and CA125, the strong survival association suggests that these tumor markers should be included in appendiceal adenocarcinoma staging similar to hCG, AFP and LDH in germ cell tumors^37^ as well as the potential future use of serial TMs measurements to track response to treatment.

Most prior studies of TM in appendiceal cancer dichotomized each TM into elevated and normal categories, however we find that treating each as a continuous variable retains important information. The distribution of values for each of CEA, CA19-9 and CA-125 were positively skewed but unimodal, so an arbitrary cutoff of the highest 10% was chosen to evaluate the survival association of highly elevated TM. We found that highly elevated CEA, CA19-9 and CA-125 were one of the strongest negative predictors of survival, similar in magnitude to stage.

In our study we comprehensively assessed the clinical utility of TMs in both operable and inoperable patients with AA and their association with tumor stage, grade^38,39^, histopathology, and mutational profile. Although metastatic tumors had higher levels of all three TM, the association of elevated TM with survival was independent of stage, gender, race, tumor grade, tumor histopathology. The analyses by tumor grade also highlighted the unique observation that patients with low-grade AA and normal CEA levels had a particular good prognosis (99% survival at 5 years and 94% survival at 5 years in metastatic disease). This observation could be explained by early diagnosis in these patients when the tumor volume is insufficient to cause elevation of the TMs, which would be associated with an especially favorable outcome. Perhaps there could be a role for treatment de-escalation in these patients, particularly considering recent prospective data showing 5-FU-based chemotherapy is ineffective in this patient population^40^.

Despite the notable distinctions between AA and CRC^2,38^, the current AJCC staging system treats them similarly^41^, lacking a hierarchical survival demonstration specific to AA and hindering clinical applicability. Our results collectively illuminate various factors in AA patients that correlate with survival and hold potential for a novel staging system for AA. As even among patients with metastatic disease, considering TMs, tumor grade, and histopathology reveals significant variations in survival outcomes.

Moreover, the study establishes a link between TMs and the mutational profile of patients with AA, demonstrating that *KRAS* and *GNAS* mutations are associated with elevated levels of both CEA and CA19-9. These findings suggest that TMs may serve as useful prognostic biomarker of disease burden, that is commonly available with low-cost, and noninvasive. Prior work in CRC has demonstrated that knockdown of mutant *KRAS* in CRC cell lines reduced CEA expression and its restoration reestablished CEA expression^42^. These findings from previous studies align with our findings and offer new insights into the possibility of identifying treatment targets for AA.

### Limitations

The study is subject to several limitations, which we would like to acknowledge. First, the retrospective design introduces inherent limitations in data collection and potential biases. As a single institution study, the findings may not be fully generalizable to other populations or healthcare settings. Moreover, the low number of patients undergoing NGS analysis (only 30% of the cohort) limits the comprehensive assessment of genetic mutations. Additionally, the selective approach in ordering tumor markers by different physicians may have led to the exclusion of certain patients from the cohort, potentially over-representing those with a more advanced disease stage. Another limitation is the lack of consideration for patients’ chemotherapy and cytoreductive surgery history. This omission is primarily due to the fact that many patients receive initial treatment at local hospitals before seeking care at our institution. Furthermore, the analysis did not investigate the differences in tumor marker levels among patients with localized disease (stage I, II, and III) due to incomplete stage data in our cohort. Despite these limitations, our study benefited from the ability to assemble a substantial cohort of 1,338 patients with AA, accompanied by comprehensive clinical data and outcomes. To the best of our knowledge, this represents the largest and most comprehensive AA patient cohort ever assembled at a single institution, providing valuable insights into the disease and its management and enabled us to control for potential variations in clinical practice, which is particularly relevant in rare diseases such as AA.

## Conclusions

In conclusion, these data demonstrate the practical value of CEA, CA19-9, and CA125 in management of AA. These biomarkers can be regarded as dependable prognostic tools for patients with AA. We suggest incorporating the measurement of these three TMs as a standard part of AA’s clinical management. Additionally, it is important to explore the potential role of TMs in AA’s tumor cell adhesion and disease progression to enhance our comprehension of the disease’s biological behavior.

## Acknowledgements

This work was supported by the Col. Daniel Connelly Memorial Fund, the National Cancer Institute (K22 CA234406 to J.P.S., Cancer Center Support Grant P30 CA016672), the Cancer Prevention & Research Institute of Texas (RR180035 to J.P.S., J.P.S. is a CPRIT Scholar in Cancer Research), and a Conquer Cancer Career Development Award (to J.P.S.). Any opinions, findings, and conclusions expressed in this material are those of the author(s) and do not necessarily reflect those of the American Society of Clinical Oncology® or Conquer Cancer. Writing support provided by Jennifer Peterson, PhD.

## Data Availability Statement

The data generated in this study are included in the supplementary tables or otherwise available upon request to the corresponding author.

