## Supplemental table 1 for "The Clinical Significance of CEA, CA19-9, and CA125 in Management of Appendiceal Adenocarcinoma"

### Supplemental Figures and Tables

#### Tables Legends

**Table S1**. Tumor marker levels and cut offs for the Main Cohort

**Table S2**. Tumor marker levels and cut offs split by tumor grade

**Table S3**. CEA level and cut off split by histopathology

**Table S4**. CA19-9 level and cut off split by histopathology

**Table S5**. CA125 level and cut off split by histopathology

**Table S6**. Number of test performed for each TM per patient

**Table S7**. Cox proportional hazards regression model A for overall survival (n=1338)

**Table S8**. Cox  proportional hazards regression model B for overall survival (n=1338)

#### Figures Legends

**Figure S1**. (A) Distribution of all patients in our cohort by year of diagnosis. (B) Distribution of all patients in our cohort by tumor histopathological grade. (C) Distribution of all patients in our cohort by tumor histopathological grade binary.

**Figure S2**. Spider plot showing % of patients tested for each tumor marker at the time of diagnosis over the last 8 years.

**Figure S3.** Violin plot showing the distribution of all patients CEA, CA19-9, and CA125 tumor markers levels split by patients sex, lines represent median levels

**Figure S4.** (A) Violin plot showing the distribution of all patients CEA tumor markers levels split by tumor histopathology, lines represent median levels. (B) Violin plot showing the distribution of all patients CA19-9 tumor markers levels split by tumor histopathology, lines represent median levels. (C) Violin plot showing the distribution of all patients CA125 tumor markers levels split by tumor histopathology, lines represent median levels.

**Figure S5.** (A) Scattered plot for patients who had CEA (on X-axis) and CA19-9 (on Y-axis) measured on the same day showing the correlation between both tumor markers. (B) Scattered plot for patients who had CEA (on X-axis) and CA125 (on Y-axis) measured on the same day showing the correlation between both tumor markers (C) Scattered plot for patients who had CA19-9 (on X-axis) and CA125 (on Y-axis) measured on the same day showing the correlation between both tumor markers. Each point represents one measurement on one day for one patient. Non parametric Spearman's correlation was used to measure the degree of association, (all $p<0.0001$)

**Figure S6.** (A) KM survival plot of patients with low grade tumor for normal, elevated, and highly elevated levels of CEA. (B) KM survival plot of patients with high grade tumor for normal, elevated, and highly elevated levels of CEA (C) KM survival plot of patients with low grade tumor for normal, elevated, and highly elevated levels of CA19-9. (D) KM survival plot of patients with high grade tumor for normal, elevated, and highly elevated levels of CA19-9. (E) KM survival plot of patients with low grade tumor for normal, elevated, and highly elevated levels of CA125. (F) KM survival plot of patients with high grade tumor for normal, elevated, and highly elevated levels of CA125.

**Figure S7.** (A) KM survival plot of all patients with normal, elevated, and highly elevated levels of CEA measured within the initial six months from the date of diagnosis. (B) KM survival plot of all patients with normal, elevated, and highly elevated levels of CA19-9 measured within the initial six months from the date of diagnosis. (C) KM survival plot of all patients with normal, elevated, and highly elevated levels of CA125 measured within the initial six months from the date of diagnosis. (D) KM survival plot of all patients with number of elevated tumor markers measured within the initial six months from the date of diagnosis. (E) Forest plot for multivariable analysis showing HR for death in the subset of patients who had their tumor markers measured within the initial six months from the date of diagnosis.

**Table S1. Tumor marker levels and cut offs for the Main Cohort**

| | CEA | CA 19-9 | CA 125 |
|---|---|---|---|
| Number of Patients (n) | 1331 | 1132 | 1165 |
| Cut off Value | > 3.8 ng/mL | > 35 U/mL | > 38 U/mL |
| Percentage Elevated (%) | 46% | 24% | 17% |
| Number Elevated | 609 | 268 | 196 |
| Highly Elevated cut off (top 10th percentile) | > 99.8 ng/mL | > 338.6 U/mL | > 99.0 U/mL |
| Percentage Highly Elevated (%) | 10% | 10% | 10% |
| Number Highly Elevated | 133 | 113 | 116 |

**Table S2. Tumor marker levels and cut offs split by tumor grade**

| | CEA | | CA 19-9 | | CA 125 | |
|---|---|---|---|---|---|---|
| Grade | Low Grade | High Grade | Low Grade | High Grade | Low Grade | High Grade |
| Number of Patients (n) | 519 | 761 | 486 | 607 | 487 | 636 |
| Cut off Value | > 3.8 ng/mL | | > 35 U/mL | | > 38 U/mL | |
| Percentage Elevated (%) | | | | | | |
| Number Elevated | 235 | 355 | 129 | 132 | 75 | 113 |
| Highly Elevated cut off (top 10th percentile) | > 99.8 ng/mL | | > 338.6 U/mL | | > 99.0 U/mL | |
| Percentage Highly Elevated (%) | 10% | | 10% | | 10% | |
| Number Highly Elevated | 53 | 78 | 41 | 70 | 37 | 76 |

**Table S3. CEA level and cut off split by histopathology**

| | CEA | | | | |
|---|---|---|---|---|---|
| Grade | Mucinous | Colonic | Goblet | Signet | Goblet & Signet |
| Number of Patients (n) | 693 | 130 | 93 | 221 | 147 |
| Cut off Value | > 3.8 ng/mL | | | | |
| Percentage Elevated (%) | 56% | 59% | 30% | 69% | 50% |
| Number Elevated | 391 | 77 | 28 | 153 | 73 |
| Mean | 64.3 | 172.8 | 12.2 | 44.6 | 10.5 |
| Std. Deviation | 244 | 593 | 62 | 126 | 28 |
| Median | 4.3 | 5.1 | 2.7 | 6.7 | 3.6 |

**Table S4. CA19-9 level and cut off split by histopathology**

| | CA19-9 | | | | |
|---|---|---|---|---|---|
| Grade | Mucinous | Colonic | Goblet | Signet | Goblet & Signet |
| Number of Patients (n) | 630 | 90 | 77 | 180 | 118 |
| Cut off Value | > 35 U/mL | | | | |
| Percentage Elevated (%) | 36% | 39% | 10% | 43% | 21% |
| Number Elevated | 224 | 35 | 8 | 77 | 25 |
| Mean | 272 | 5829 | 35 | 408 | 72 |
| Std. Deviation | 1431 | 37295 | 130 | 1413 | 390 |
| Median | 20 | 21 | 13 | 27 | 14 |

**Table S5. CA125 level and cut off split by histopathology**

| | CA125 | | | | |
|---|---|---|---|---|---|
| **Grade** | **Mucinous** | **Colonic** | **Goblet** | **Signet** | **Goblet & Signet** |
| **Number of Patients (n)** | **631** | **84** | **79** | **190** | **130** |
| **Cut off Value** | **> 38 U/mL** | | | | |
| **Percentage Elevated (%)** | **25%** | **25%** | **13%** | **38%** | **26%** |
| **Number Elevated** | **157** | **21** | **10** | **73** | **34** |
| **Mean** | **34.0** | **59.0** | **21.5** | **59.2** | **44.5** |
| **Std. Deviation** | **49.6** | **168.4** | **26.7** | **89.8** | **74.5** |
| **Median** | **15.7** | **15.4** | **12.9** | **21.6** | **17.7** |

**Table S6. Number of test performed for each TM per patient**

| | CEA | CA 19-9 | CA 125 |
|---|---|---|---|
| **Mode** | **1** | **1** | **1** |
| **Approximate Median** | **4** | **3** | **3** |
| **Approximate Mean** | **7** | **5** | **5** |
| **Min** | **1** | **1** | **1** |
| **Max** | **98** | **60** | **36** |

**Table S7. Cox proportional hazards regression model A for overall survival (n=1338)**

| Clinical Characteristics | Univariate Analysis | | | | Multivariate Analysis | | | |
|---|---|---|---|---|---|---|---|---|
| | HR | P value | 95 CI% | | HR | P value | 95 CI% | |
| Male gender | | reference | | | | reference | | |
| Female gender | 0.6589 | 0.0036 | 0.4977 | 0.8722 | 0.78 | 0.16 | 0.54 | 1.1 |
| Race white | | reference | | | | Reference | | |
| Race Black or African American | 1.9 | 0.0072 | 1.19 | 3.033 | 2.9 | 0.0005 | 1.6 | 5.1 |
| Race Hispanic/ Latino | 1.264 | 0.3156 | 0.7999 | 1.997 | 2.5 | 0.0017 | 1.4 | 4.5 |
| Race Asian | 1.531 | 0.2717 | 0.7164 | 3.272 | | | | |
| Race Others | 0.7099 | 0.6305 | 0.1758 | 2.867 | | | | |
| Smoking status Never | | reference | | | | reference | | |
| Smoking status Former | 1.294 | 0.1215 | 0.9338 | 1.794 | | | | |
| Smoking status Smoker | 0.9791 | 0.9566 | 0.4575 | 2.095 | | | | |
| Alcohol use status Never | | reference | | | | reference | | |
| Alcohol use status Yes | 0.7444 | 0.0462 | 0.5569 | 0.995 | 1.2 | 0.45 | 0.8 | 1.7 |
| Age at diagnosis | 1.015 | 0.0218 | 1.002 | 1.028 | 1 | 0.28 | 0.99 | 1 |
| Tumor Histopathology Mucinous | | reference | | | | reference | | |
| Tumor Histopathology Colonic | 2.855 | 9.32E-06 | 1.795 | 4.539 | 1.9 | 0.063 | 0.97 | 3.7 |
| Tumor Histopathology Goblet | 0.2172 | 0.1297 | 0.03014 | 1.565 | | | | |
| Tumor Histopathology Signet | 4.278 | 5.24E-17 | 3.045 | 6.009 | 3.1 | 1.80E-07 | 2 | 4.8 |
| Tumor Histopathology Goblet/Signet | 3.228 | 6.13E-08 | 2.112 | 4.933 | 3.3 | 0.0013 | 1.9 | 5.8 |
| Low Grade tumor | | reference | | | | reference | | |
| High Grade Tumor | 3.782 | 1.61E-14 | 2.693 | 5.31 | 3.5 | 1.70E-08 | 2.3 | 5.4 |
| Normal CEA | | reference | | | | reference | | |
| Elevated CEA | 4.997 | 6.52E-16 | 3.382 | 7.382 | 2.8 | 0.0001 | 1.7 | 4.9 |
| Normal CA19-9 | | reference | | | | reference | | |
| Elevated CA19-9 | 3.088 | 1.16E-11 | 2.23 | 4.277 | 1.5 | 0.028 | 1 | 2.2 |
| Normal CA125 | | reference | | | | reference | | |
| Elevated CA125 | 5.936 | 2.97E-27 | 4.298 | 8.198 | 3.2 | 1.70E-09 | 2.2 | 4.7 |
| Localized Disease | | reference | | | | reference | | |
| Metastatic Disease | 11.51 | 1.33E-6 | 4.275 | 30.99 | 9.8 | 0.0017 | 2.4 | 41 |

**Table S8. Cox proportional hazards regression model B for overall survival (n=1338)**

| | Univariate Analysis | | | | Multivariate Analysis | | | |
|---|---|---|---|---|---|---|---|---|
| Clinical Characteristics | HR | P value | 95 CI% | | HR | P value | 95 CI% | |
| Male gender | reference | | | | reference | | | |
| Female gender | 0.6589 | 0.0036 | 0.4977 | 0.8722 | 0.69 | 0.017 | 0.51 | 0.94 |
| Race white | reference | | | | reference | | | |
| Race Black or African American | 1.9 | 0.0072 | 1.19 | 3.033 | 1.9 | 0.013 | 1.1 | 3.2 |
| Race Hispanic/ Latino | 1.264 | 0.3156 | 0.7999 | 1.997 | 1.9 | 0.013 | 1.1 | 3.1 |
| Race Asian | 1.531 | 0.2717 | 0.7164 | 3.272 | | | | |
| Race Others | 0.7099 | 0.6305 | 0.1758 | 2.867 | | | | |
| Smoking status Never | reference | | | | reference | | | |
| Smoking status Former | 1.294 | 0.1215 | 0.9338 | 1.794 | | | | |
| Smoking status Smoker | 0.9791 | 0.9566 | 0.4575 | 2.095 | | | | |
| Alcohol use status Never | reference | | | | reference | | | |
| Alcohol use status Yes | 0.7444 | 0.0462 | 0.5569 | 0.995 | 1 | 0.99 | 0.73 | 1.4 |
| Age at diagnosis | 1.016 | 0.0127 | 1.003 | 1.029 | 1 | 0.5 | 0.99 | 1 |
| Tumor Histopathology Mucinous | reference | | | | reference | | | |
| Tumor Histopathology Colonic | 2.855 | 9.32E-06 | 1.795 | 4.539 | 2 | 0.0071 | 1.2 | 3.4 |
| Tumor Histopathology Goblet | 0.2172 | 0.1297 | 0.03014 | 1.565 | | | | |
| Tumor Histopathology Signet | 4.278 | 5.24E-17 | 3.045 | 6.009 | 3 | 7.60E-09 | 2.1 | 4.4 |
| Tumor Histopathology Goblet/Signet | 3.228 | 6.13E-08 | 2.112 | 4.933 | 3.1 | 6.10E-06 | 1.9 | 5 |
| Low Grade tumor | reference | | | | reference | | | |
| High Grade Tumor | 3.782 | 1.61E-14 | 2.693 | 5.31 | 3.4 | 1.10E-09 | 2.3 | 5 |
| Normal level of all the 3 TMs | reference | | | | reference | | | |
| Elevated one TM | 6.831 | 5.13E-11 | 3.85 | 12.12 | 4 | 1.70E-07 | 2.1 | 7.7 |
| Elevated two TM | 10.14 | 3.31E-15 | 5.698 | 18.04 | 6.4 | 2.90E-08 | 3.3 | 12 |
| Elevated three TM | 15.97 | 3.30E-19 | 8.709 | 29.27 | 11 | 1.00E-11 | 5.4 | 21 |
| Localized Disease | reference | | | | reference | | | |
| Metastatic Disease | 11.51 | 1.33E-06 | 4.275 | 30.99 | 9.3 | 0.0002 | 2.9 | 30 |

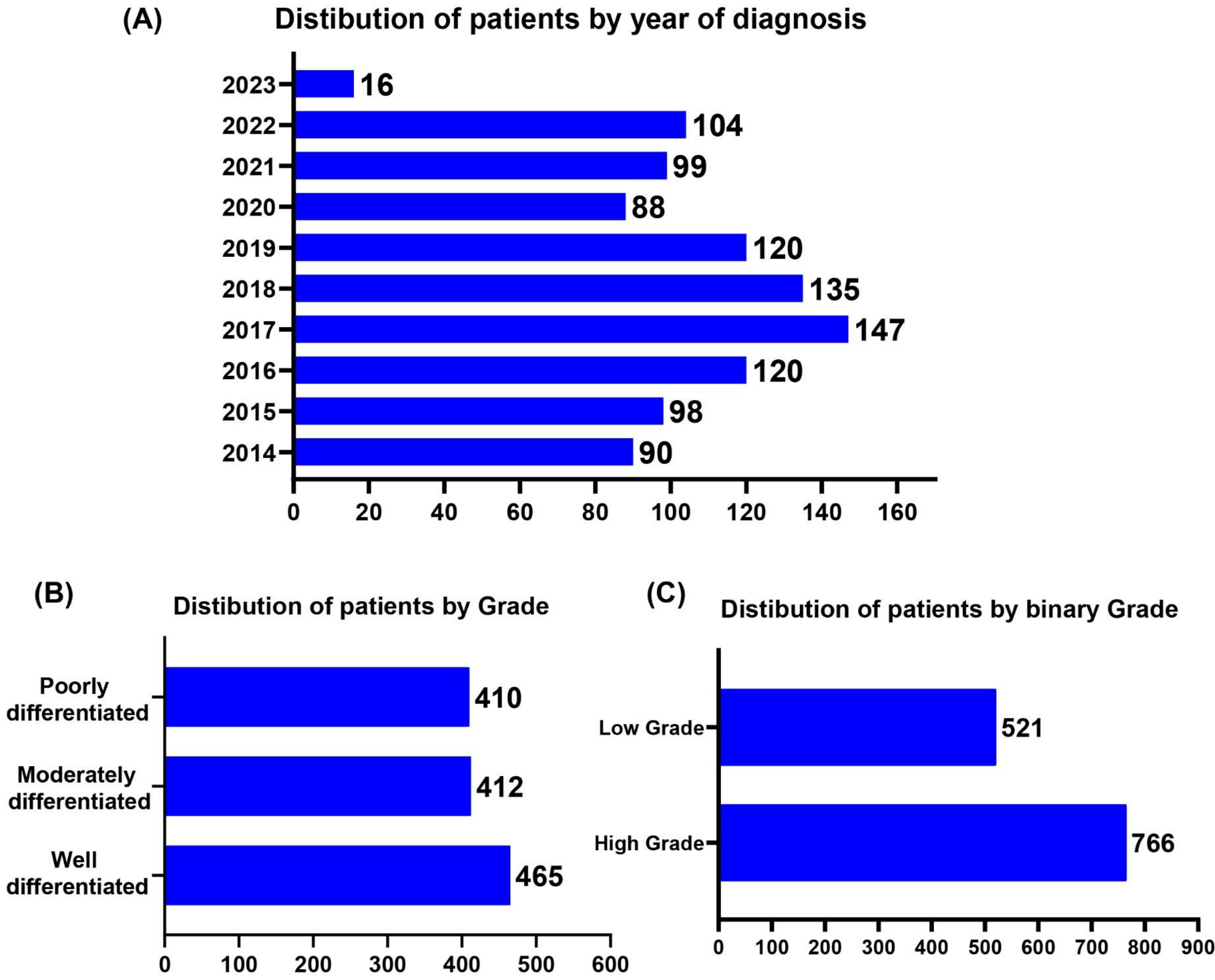
(A)
Distibution of patients by year of diagnosis
2023
2022
2021
2020
2019
2018
2017
2016
2015
2014
16
104
99
88
120
135
147
120
98
90
0
20
40
60
80
100
120
140
160
(B)
Distibution of patients by Grade
Poorly differentiated
Moderately differentiated
Well differentiated
410
412
465
0
100
200
300
400
500
600
(C)
Distibution of patients by binary Grade
Low Grade
High Grade
521
766
0
100
200
300
400
500
600
700
800
900

Fig S1

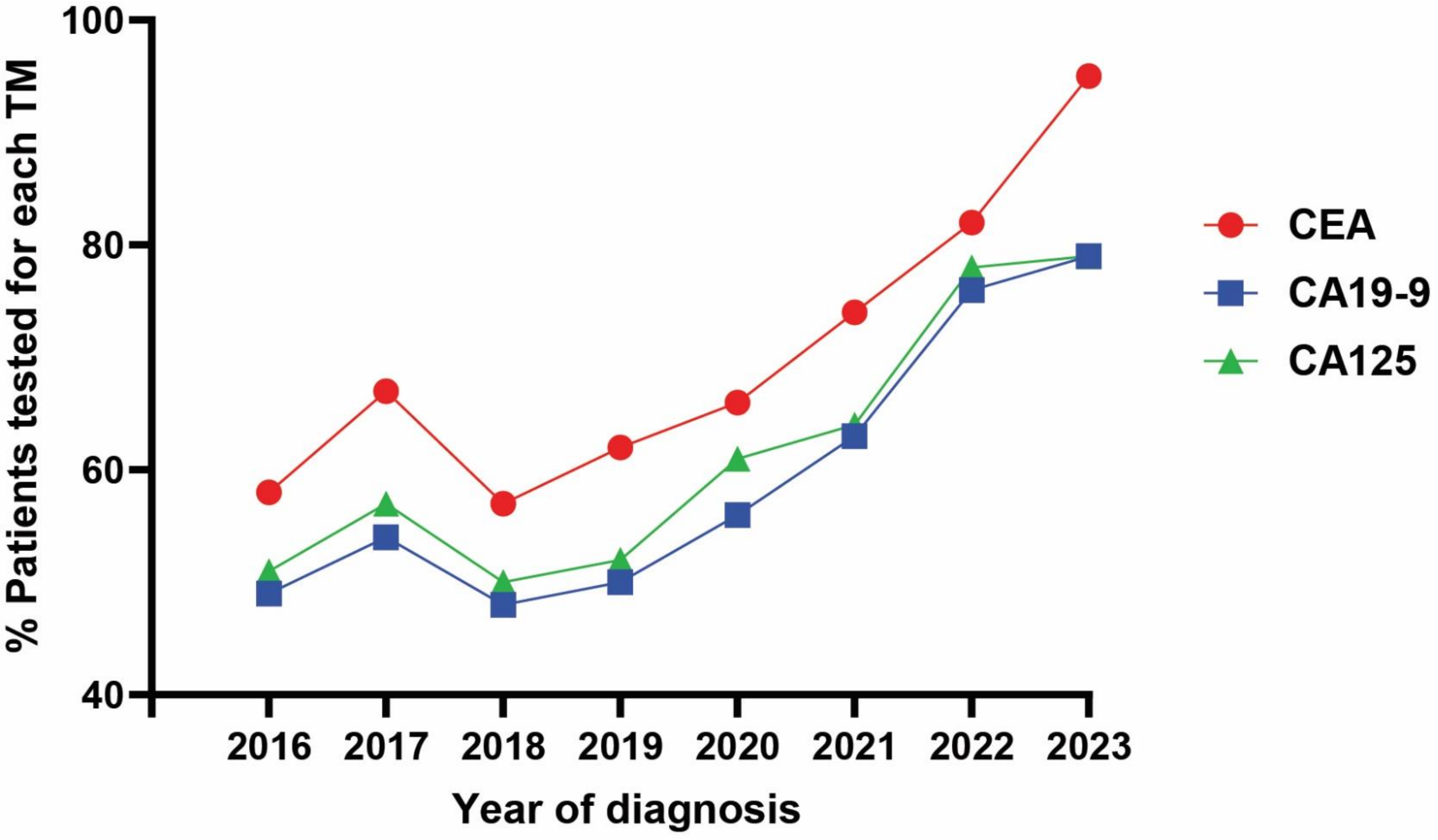

% Patients tested for each TM
100
80
60
40
2016
2017
2018
2019
2020
2021
2022
2023
Year of diagnosis
CEA
CA19-9
CA125


Fig S2

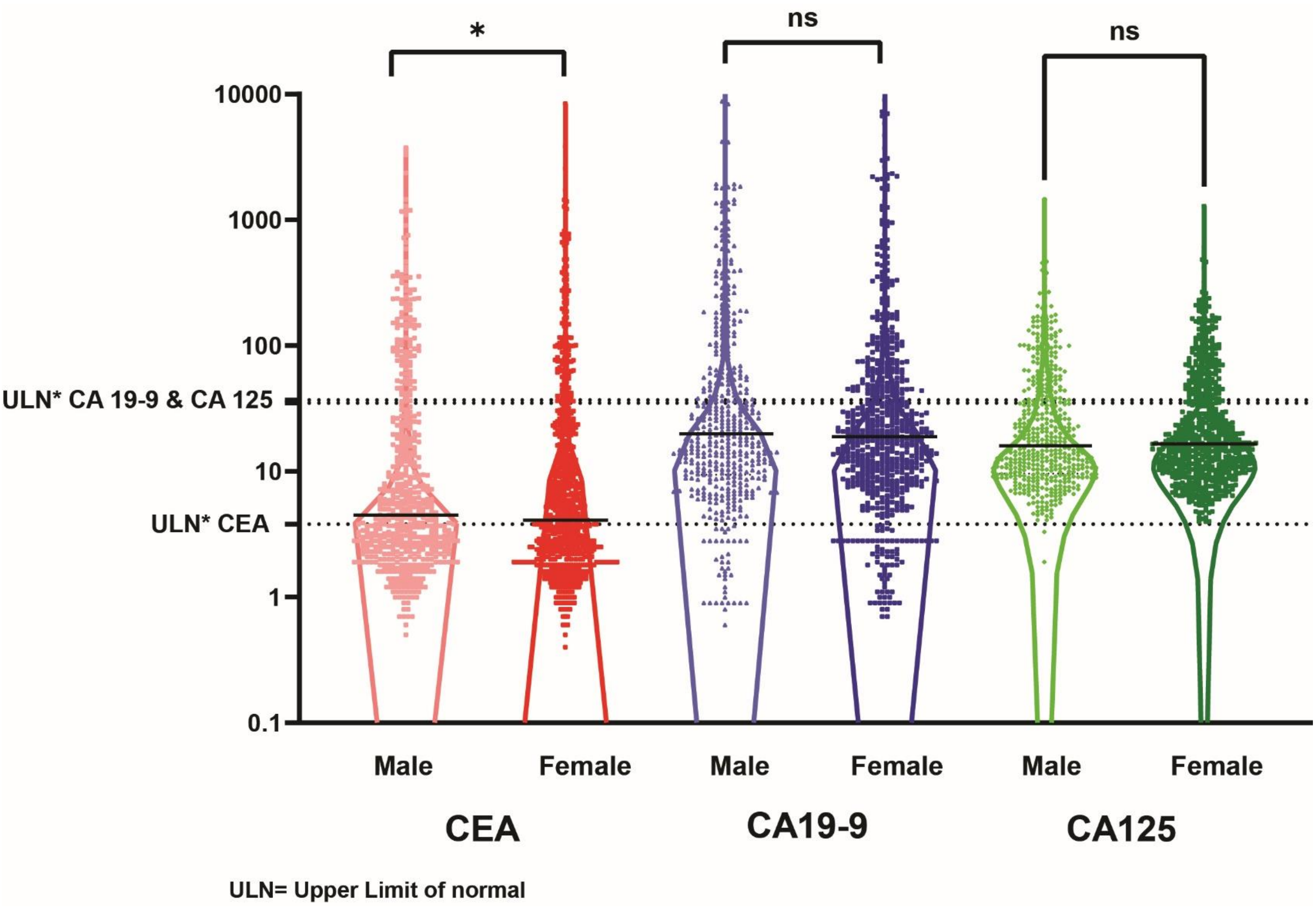
*
ns
ns
10000
1000
100
ULN* CA 19-9 & CA 125
10
ULN* CEA
1
0.1
Male
Female
Male
Female
Male
Female
CEA
CA19-9
CA125
ULN= Upper Limit of normal

**Fig S3**

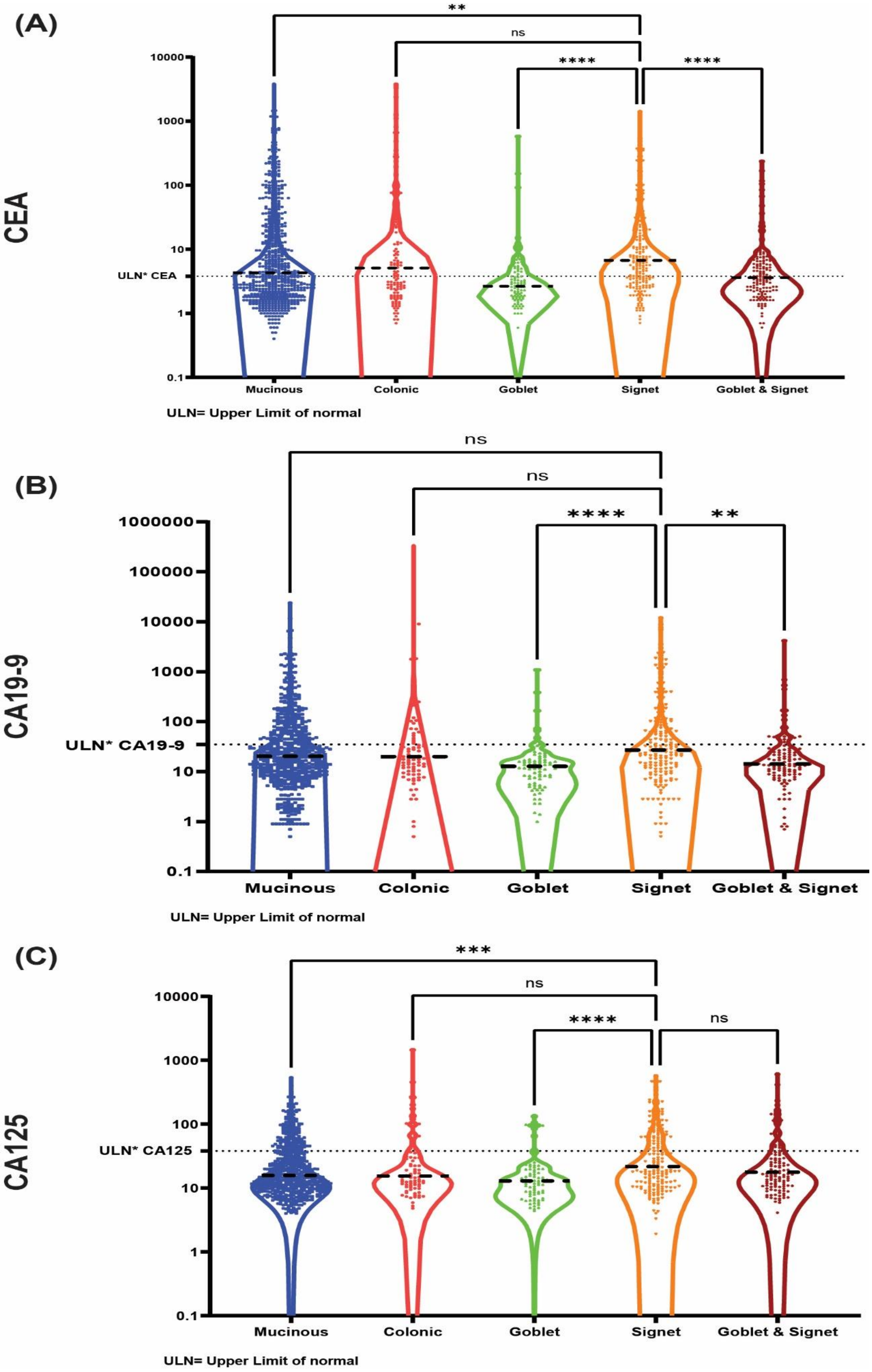
(A)
CEA
ULN* CEA
10000
1000
100
10
1
0.1
**
ns
****
****
Mucinous
Colonic
Goblet
Signet
Goblet & Signet
ULN= Upper Limit of normal
(B)
CA19-9
ULN* CA19-9
1000000
100000
10000
1000
100
10
1
0.1
ns
ns
****
**
Mucinous
Colonic
Goblet
Signet
Goblet & Signet
ULN= Upper Limit of normal
(C)
CA125
ULN* CA125
10000
1000
100
10
1
0.1
***
ns
****
ns
Mucinous
Colonic
Goblet
Signet
Goblet & Signet
ULN= Upper Limit of normal



**Fig S4**

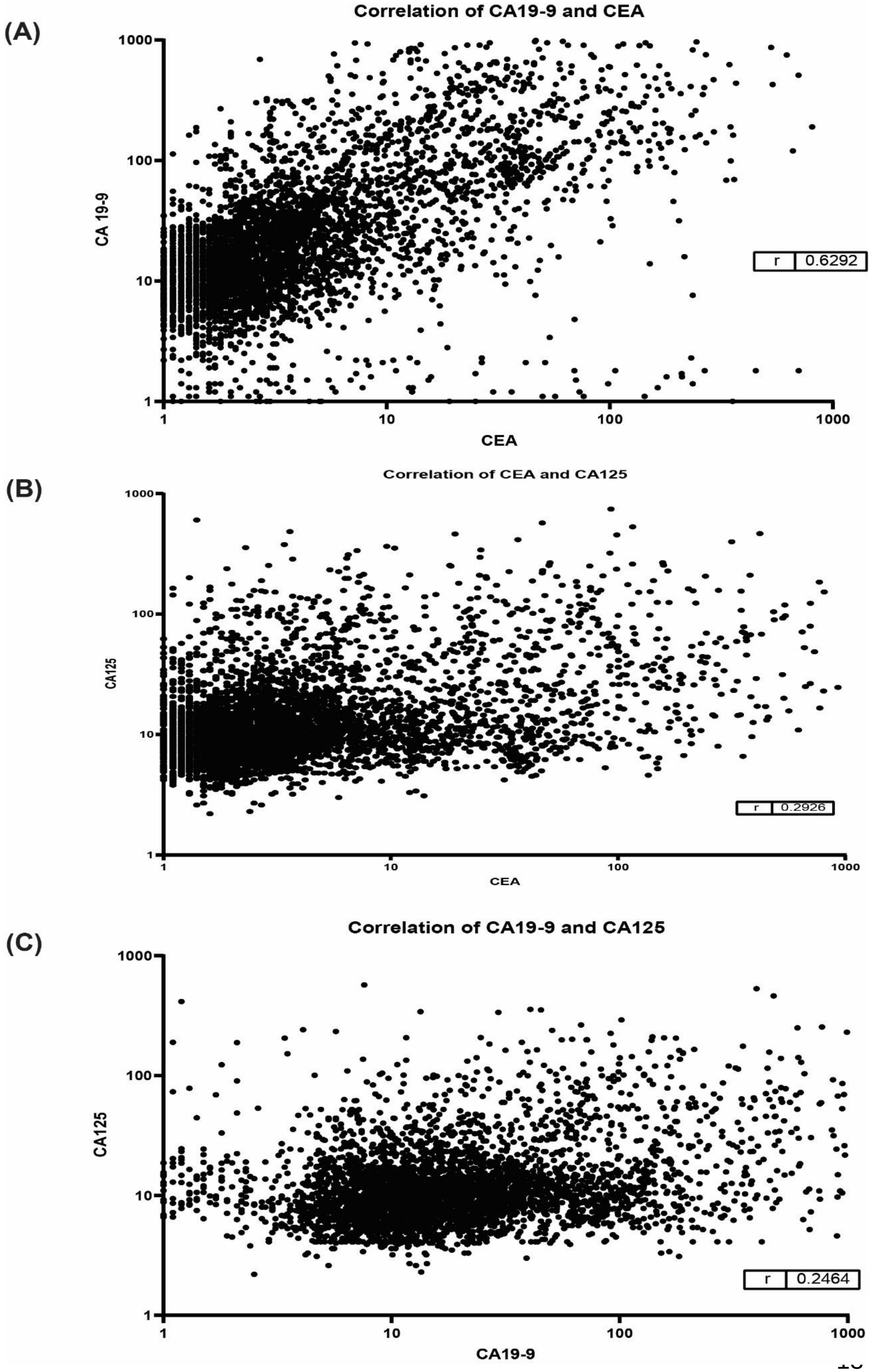
(A)
Correlation of CA19-9 and CEA
CA 19-9
CEA
1000
100
10
1
r 0.6292
(B)
Correlation of CEA and CA125
CA125
CEA
r 0.2926
(C)
Correlation of CA19-9 and CA125
CA125
CA19-9
r 0.2464

**Fig S5**

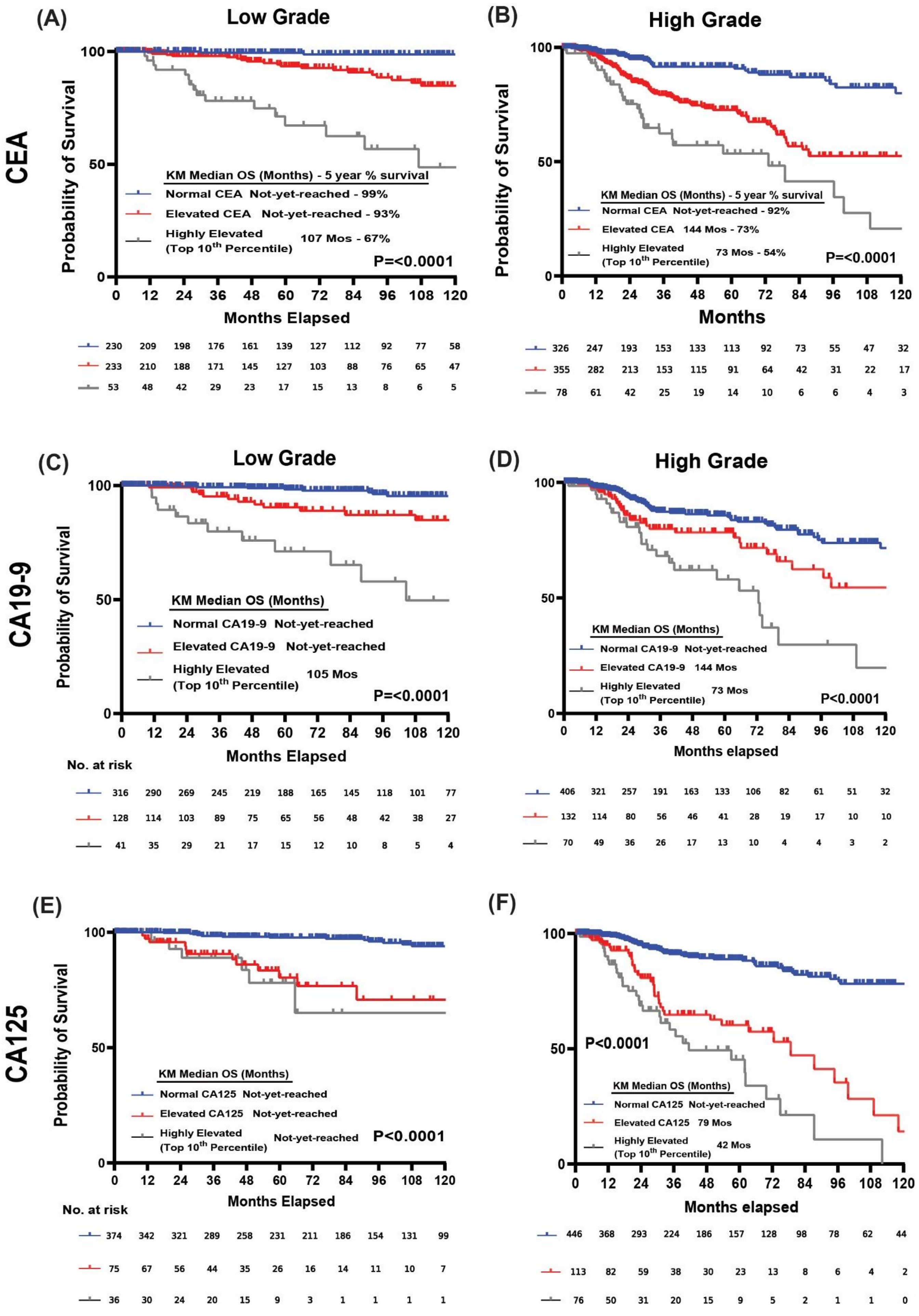

(A)
Low Grade
CEA
Probability of Survival
KM Median OS (Months) - 5 year % survival
Normal CEA Not-yet-reached - 99%
Elevated CEA Not-yet-reached - 93%
Highly Elevated (Top 10th Percentile) 107 Mos - 67%
P=<0.0001
Months Elapsed
230 209 198 176 161 139 127 112 92 77 58
233 210 188 171 145 127 103 88 76 65 47
53 48 42 29 23 17 15 13 8 6 5
(B)
High Grade
Probability of Survival
KM Median OS (Months) - 5 year % survival
Normal CEA Not-yet-reached - 92%
Elevated CEA 144 Mos - 73%
Highly Elevated (Top 10th Percentile) 73 Mos - 54%
P=<0.0001
Months
326 247 193 153 133 113 92 73 55 47 32
355 282 213 153 115 91 64 42 31 22 17
78 61 42 25 19 14 10 6 6 4 3
(C)
Low Grade
CA19-9
Probability of Survival
KM Median OS (Months)
Normal CA19-9 Not-yet-reached
Elevated CA19-9 Not-yet-reached
Highly Elevated (Top 10th Percentile) 105 Mos
P=<0.0001
Months Elapsed
No. at risk
316 290 269 245 219 188 165 145 118 101 77
128 114 103 89 75 65 56 48 42 38 27
41 35 29 21 17 15 12 10 8 5 4
(D)
High Grade
KM Median OS (Months)
Normal CA19-9 Not-yet-reached
Elevated CA19-9 144 Mos
Highly Elevated (Top 10th Percentile) 73 Mos
P<0.0001
Months elapsed
406 321 257 191 163 133 106 82 61 51 32
132 114 80 56 46 41 28 19 17 10 10
70 49 36 26 17 13 10 4 4 3 2
(E)
CA125
Probability of Survival
KM Median OS (Months)
Normal CA125 Not-yet-reached
Elevated CA125 Not-yet-reached
Highly Elevated (Top 10th Percentile) Not-yet-reached
P<0.0001
Months Elapsed
No. at risk
374 342 321 289 258 231 211 186 154 131 99
75 67 56 44 35 26 16 14 11 10 7
36 30 24 20 15 9 3 1 1 1 1
(F)
P<0.0001
KM Median OS (Months)
Normal CA125 Not-yet-reached
Elevated CA125 79 Mos
Highly Elevated (Top 10th Percentile) 42 Mos
Months elapsed
446 368 293 224 186 157 128 98 78 62 44
113 82 59 38 30 23 13 8 6 4 2
76 50 31 20 15 9 5 2 1 1 0


**Fig S6**

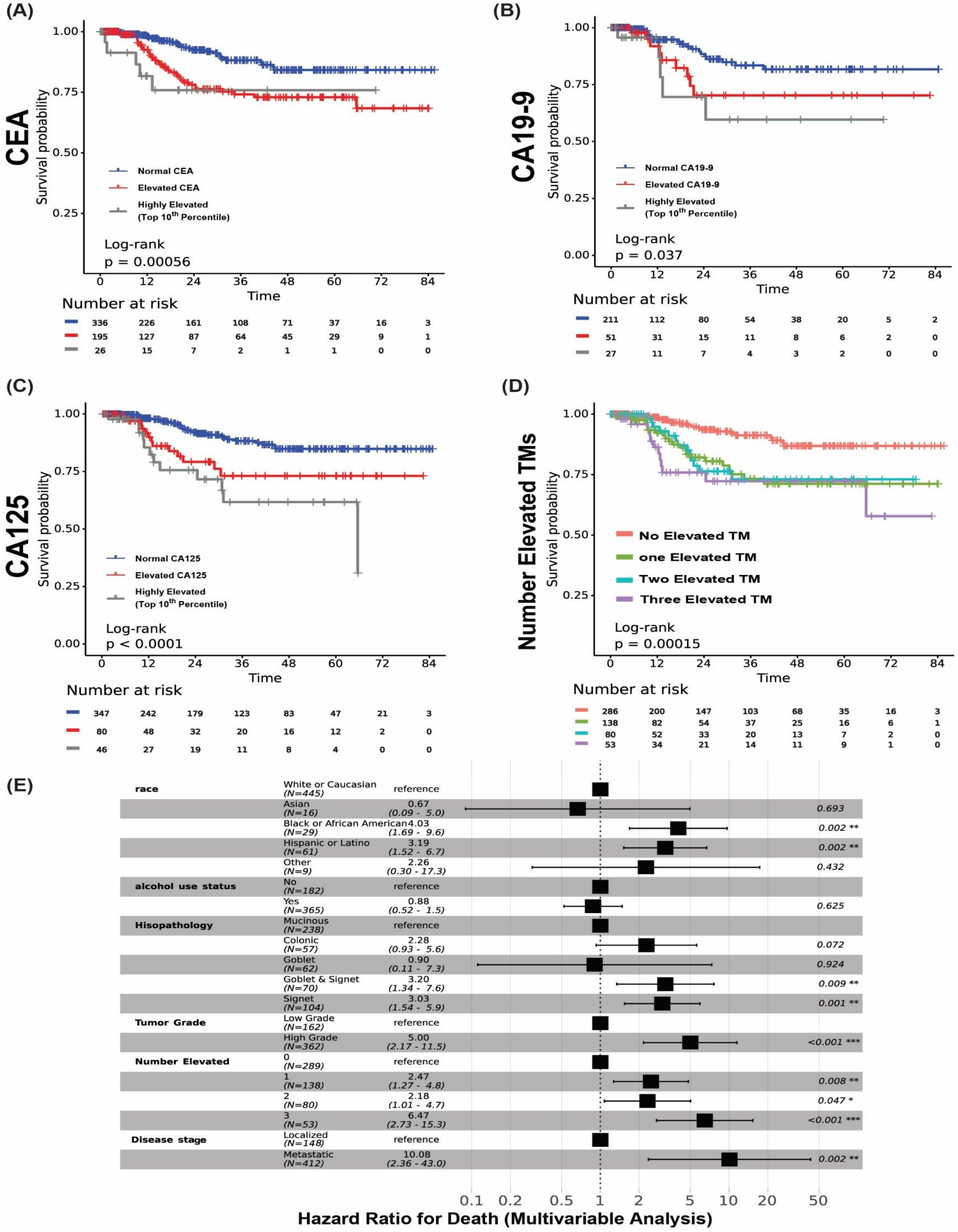
(A)
CEA
Survival probability
Normal CEA
Elevated CEA
Highly Elevated
(Top 10th Percentile)
Log-rank
p = 0.00056
Time
Number at risk
336 226 161 108 71 37 16 3
195 127 87 64 45 29 9 1
26 15 7 2 1 1 0 0
(B)
CA19-9
Survival probability
Normal CA19-9
Elevated CA19-9
Highly Elevated
(Top 10th Percentile)
Log-rank
p = 0.037
Time
Number at risk
211 112 80 54 38 20 5 2
51 31 15 11 8 6 2 0
27 11 7 4 3 2 0 0
(C)
CA125
Survival probability
Normal CA125
Elevated CA125
Highly Elevated
(Top 10th Percentile)
Log-rank
p < 0.0001
Time
Number at risk
347 242 179 123 83 47 21 3
80 48 32 20 16 12 2 0
46 27 19 11 8 4 0 0
(D)
Number Elevated TMs
Survival probability
No Elevated TM
one Elevated TM
Two Elevated TM
Three Elevated TM
Log-rank
p = 0.00015
Time
Number at risk
286 200 147 103 68 35 16 3
138 82 54 37 25 16 6 1
80 52 33 20 13 7 2 0
53 34 21 14 11 9 1 0
(E)
race
White or Caucasian (N=445) reference
Asian (N=16) 0.67 (0.09 - 5.0) 0.693
Black or African American (N=29) 4.03 (1.69 - 9.6) 0.002 **
Hispanic or Latino (N=61) 3.19 (1.52 - 6.7) 0.002 **
Other (N=9) 2.26 (0.30 - 17.3) 0.432
alcohol use status
No (N=182) reference
Yes (N=365) 0.88 (0.52 - 1.5) 0.625
Hisopathology
Mucinous (N=238) reference
Colonic (N=57) 2.28 (0.93 - 5.6) 0.072
Goblet (N=62) 0.90 (0.11 - 7.3) 0.924
Goblet & Signet (N=70) 3.20 (1.34 - 7.6) 0.009 **
Signet (N=104) 3.03 (1.54 - 5.9) 0.001 **
Tumor Grade
Low Grade (N=162) reference
High Grade (N=362) 5.00 (2.17 - 11.5) <0.001 ***
Number Elevated
0 (N=289) reference
1 (N=138) 2.47 (1.27 - 4.8) 0.008 **
2 (N=80) 2.18 (1.01 - 4.7) 0.047 *
3 (N=53) 6.47 (2.73 - 15.3) <0.001 ***
Disease stage
Localized (N=148) reference
Metastatic (N=412) 10.08 (2.36 - 43.0) 0.002 **
Hazard Ratio for Death (Multivariable Analysis)

**Fig S7**
